# Psychiatric morbidity among patients living with epilepsy at a tertiary referral hospital in Western Kenya: A cross-sectional study

**DOI:** 10.64898/2026.07.11.26357815

**Authors:** Agnes Achieng Odhiambo, Daniel Waiganjo.C Kinyanjui, Robina Kerubo Momanyi

**Author notes:** Corresponding author: (AAO).

## Abstract

**Background:** Psychiatric comorbidities commonly have a negative impact on epilepsy outcomes. However, they are continuously ignored in routine epilepsy care, with focus directed more towards seizure control. There is paucity of data on the burden of psychiatric morbidity among those living with epilepsy in Kenya. This study sought to determine the prevalence and associated factors of psychiatric morbidity among patients living with epilepsy at a tertiary referral hospital in Western Kenya.

**Methods:** This was a descriptive cross-sectional study. Consecutive sampling was used to recruit participants, with a sample size of 278. Data were collected using a structured pretested sociodemographic and clinical characteristics questionnaire, and the Mini International Neuropsychiatric Interview (MINI), and analyzed using STATA version 16. Pearson Chi-square test/Fisher’s Exact test and logistic regression were used to assess relationships at bivariate and multivariate levels respectively.

**Results:** The prevalence of psychiatric morbidity was 52.2%. Major depressive disorder was the most prevalent (36%), followed by anxiety disorders (26.2%), psychotic disorders (16.9%), and suicidality (15.1%). Casual/self-employment (aOR=2.590, p=0.020), seizure-related physical trauma (aOR=4.032, p=0.004), antiepileptic polytherapy (aOR=4.280, p=0.001), frequent seizures (aOR=3.801, p<0.001), and comorbid medical conditions (aOR=5.478, p=0.047) were independent predictors of psychiatric morbidity. Having attained a tertiary level of education was protective against psychiatric morbidity (aOR=0.221, p=0.036).

**Conclusion:** More than half of the patients living with epilepsy had at least one psychiatric comorbidity. Routine psychiatric screening and integration of mental health services in epilepsy care is essential to improve clinical outcomes.

## Introduction

Epilepsy is a chronic neurological disorder characterized by recurrent unprovoked seizures resulting from abnormal electrical activity in the brain. It affects over 50 million people worldwide, and remains one of the most common neurological disorders globally; burdening more people in low-and middle-income countries [1]. Epilepsy impact extends beyond the seizures, as there is an increasing awareness of its significant psychiatric and psychosocial consequences [2–4].

Psychiatric morbidity is prevalent among individuals living with epilepsy, yet underappreciated in general clinical practice [5]. Numerous studies have demonstrated a significantly increased risk of psychiatric comorbidity in individuals living with epilepsy relative to the general population [6–8], notably depression and anxiety disorders [9–13]. It is estimated that as many as 1 in 3 individuals living with epilepsy will have a psychiatric disorder at some point during their lifetime [14], with higher rates noted in those with refractory epilepsy [15].

The co-occurrence of epilepsy and psychiatric illness is multifaceted and reciprocal, sharing common neurobiological mechanisms of limbic system dysfunction, neurotransmitter abnormalities, and structural brain abnormalities involving regions that are also responsible for mood and behavior regulation [16,17]. Psychosocial factors also play a role, with the effects of social stigma, unemployment and social isolation leading to diminished quality of life and contributing to psychological problems [15–19]. Factors such as seizure frequency, antiepileptic polytherapy, physical trauma from seizures, and comorbid medical conditions have been linked to psychiatric symptoms among individuals living with epilepsy [20–25].

Despite the established association, psychiatric comorbidities in epilepsy remain significantly underdiagnosed and undertreated, especially in resource constrained regions [26]. In Sub-Saharan Africa, including Kenya, mental health screening is hardly ever incorporated into routine epilepsy management. This results in a substantial treatment gap and poor clinical outcomes.

Moi Teaching and Referral Hospital (MTRH) is a large tertiary hospital in western Kenya that serves a broad geographical region, and offers specialized neurological care for patients living with epilepsy. However, there is paucity of locally derived data on the prevalence and correlates of psychiatric impairment in this population. This information is essential for informing strategies for improved care and integration of mental health services in routine epilepsy care.

This study sought to estimate the prevalence of psychiatric morbidity and identify factors associated with it among patients living with epilepsy on follow up at Moi Teaching and Referral Hospital. These objectives were achieved through the generation of evidence on the burden and determinants of psychiatric morbidity among individuals living with epilepsy in this setting.

## Methods

### Study design

This was a descriptive cross-sectional study design conducted between February 2024 and January 2025.

### Study setting

The study took place at the adult neurology outpatient clinic of Moi Teaching and Referral Hospital (MTRH) in Eldoret City, Uasin Gishu County; situated 320 km north-west of Nairobi, the capital city of Kenya. It is the second largest public referral hospital in the country, and serves a large and diverse population within the western Kenya region and beyond. It offers several specialized services with the outpatient neurology clinic providing follow up care to patients with neurological conditions, including epilepsy.

### Study population

The study population consisted of adult patients living with epilepsy attending the weekly neurology clinic at MTRH. About 32 patients visit the clinic every Thursday of the week; which totals to approximately 128 patients in a month. From the clinic register, 35% of the attending patients have epilepsy; this accounts to approximately 45 patients living with epilepsy per month; and 11 per week on average.

### Sample Size

Sample size was calculated using Fisher’s formula. A previous similar study [27] had reported a prevalence of 79.2% for psychiatric morbidity in epilepsy (p=0.792). Thus, with a confidence interval of 95% (Z=1.96), and a precision of 5% (d=0.05), the minimum sample size was 253. Accounting for an attrition rate of 10%, the final sample size was 278. Consecutive sampling was employed, whereby all eligible patients were recruited until the required sample size was attained.

### Eligibility criteria

All adult patients with a confirmed diagnosis of epilepsy who were willing to participate in the study and had the capacity to provide informed consent were included. Patients who scored less than 15 on the University of California, San Diego Brief Assessment of Capacity to Consent (UBACC) tool were excluded from the study as they were considered to be lacking the capacity to provide informed consent.

### Data collection

Data were collected from February 2024 to January 2025 using interviewer-administered instruments. Capacity to consent was determined using the UBACC tool.

A structured pretested questionnaire was used to record sociodemographic and clinical data, including age, sex, marital status, educational level, employment status, residence, religion, duration of epilepsy, frequency of seizures, number of antiepileptic drugs, seizure related physical trauma, family history of epilepsy/mental illness and comorbid medical illnesses (see S1 File).

The Mini International Neuropsychiatric Interview (MINI) version 7.0 was used to measure the prevalence of various psychiatric disorders. MINI is a validated and structured diagnostic interview administered by trained interviewers, and it screens for a range of psychiatric diagnoses including depression, anxiety disorders, psychosis, suicidality, bipolar disorder, post-traumatic stress disorder and substance use disorders. Interviews were done in a private room for purposes of confidentiality. Participants who had psychiatric diagnoses were linked to appropriate mental health services.

### Data quality assurance

The researcher-designed questionnaire was pretested on a group of patients living with epilepsy already linked to the mental health clinic to determine clarity and practicability. The researcher conducted all the interviews. Daily data checking was done for completeness and accuracy. Data was entered into a password protected database accessible only to the researcher and statistician.

### Data management and analysis

Data were entered, cleaned, and analyzed using Stata V.16.0 (StataCorp LLC, College Station, TX, USA). During data analysis, variables such as age, marital status and employment status were regrouped due to smaller sample sizes in some categories leading to wide confidence intervals. This was carefully done based on shared characteristics, while ensuring that the resulting categories remained meaningful and interpretable. Categorical variables were summarized with frequency and percentages, and continuous variables were summarized with mean/median and standard deviation/interquartile range.

The dependent/outcome variable was psychiatric morbidity. The associations were examined by Chi-square test or Fisher’s exact test for categorical variables, and independent t-test or Mann-Whitney U test for continuous variables. Variables with p value of < 0.2 at bivariate analysis were further examined in the multivariate logistic regression model to determine independent predictors of psychiatric morbidity. The adjusted odds ratio with 95% confidence interval was reported, and statistical significance was considered if p value was < 0.05. The results were presented using tables and figures. The dataset is provided as supporting information (see S2 File).

### Ethical considerations

Ethical approval was granted by the Moi University/MTRH Institutional Research and Ethics Committee (IREC) (Approval No. 0004617), and the National Commission for Science, Technology and Innovation (NACOSTI) (License No. NACOSTI/P/24/32716). In addition, permission was sought and obtained from the hospital management.

All respondents gave written informed consent prior to the interview. All responses were treated with high level of confidentiality. Participants were given unique identification numbers. Data was securely kept, and was used solely for research purposes. Participants with psychiatric diagnoses were linked to appropriate mental health services. The ethical approval letter and NACOSTI research authorization permit are provided as supporting information (see S3 File and S4 File).

## Results

309 potential participants were approached and requested to participate in the study. 17 declined to participate in the study due to lack of interest to be part of the study and were respectfully left out. Of the 292 who were willing to participate, 14 lacked capacities to consent and were therefore excluded. The remaining 278 were recruited into the study after they had provided written informed consent. There was no missing data.

### Sociodemographic characteristics of study participants

There was a total of 278 participants, giving a response rate of 100%. The ages of participants ranged from 18 to 67years (mean=30.3, ± 9). Majority of the participants (n= 197, 70.9%) were between 18 and 34 years of age. Gender was almost equally distributed (n=144, 51.8% males, n=134, 48.2% females). Slightly more than half of the participants (n=169, 60.8%) were single. Few (n=40, 14.4%) participants had only primary level of education. Almost half of the participants (n=122,43.9%) were casually/self-employed, or unemployed (n=121,43.5%). Slightly more participants lived in urban areas (n=149, 53.6%), and majority were Christians (n=274, 98.6%). Findings are as illustrated in Table 1.

**Table 1.** Sociodemographic characteristics of the study participants.

| Variable | Category | Frequency (n) | Percentage (%) |
| --- | --- | --- | --- |
| Age in years | Mean (SD) | 30.3 (9.0) |  |
|  | Range | 18-67 |  |
| Age in years<br>(age brackets) | 18-34 | 197 | 70.9 |
|  | 35-49 | 73 | 26.2 |
|  | >50 | 8 | 2.9 |
| Gender | Male | 144 | 51.8 |
|  | Female | 134 | 48.2 |
| Marital status | Married | 82 | 29.5 |
|  | Single | 169 | 60.8 |
|  | Separated/Widowed | 27 | 9.7 |
| Education level | Primary | 40 | 14.4 |
|  | Secondary | 159 | 57.2 |
|  | Tertiary | 79 | 28.4 |
| Employment status | Formally employed | 35 | 12.6 |
|  | Casually/Self employed | 122 | 43.9 |
|  | Unemployed | 121 | 43.5 |
| Residence | Rural | 129 | 46.4 |
|  | Urban | 149 | 53.6 |
| Religion | Christian | 274 | 98.6 |
|  | Muslim | 4 | 1.4 |

### Clinical characteristics of the study participants

The duration of living with epilepsy ranged from 1 to 29 years, with a median of 5 years (IQR: 3.0-8.0). Majority of the participants (n=226, 81.3%) had lived with epilepsy for ≤ 10 years. A third (n=83, 29.9%) of the respondents had sustained bodily injuries secondary to seizures. 37.8 % (n=105) of the participants had experienced at least one seizure in the preceding month, while 66.9% (n=186) were on antiepileptic monotherapy. Of the participants, 4.7% (n=13) reported a family history of epilepsy, 9.7% (27) had a family history of mental illness and 11.9% (n=33) had comorbid medical conditions. The findings are displayed in Table 2.

**Table 2.** Clinical Characteristics of the study participants.

| Variable | Category | Frequency (n) | Percentage (%) |
| --- | --- | --- | --- |
| Duration of living with epilepsy | Median (IQR) | 5.0 (3.0-8.0) |  |
|  | Range | 1-29 |  |
|  | ≤ 10 years | 226 | 81.3 |
|  | ≥11 years | 52 | 18.7 |
| Seizure related physical trauma | No | 195 | 70.1 |
|  | Yes | 83 | 29.9 |
| Number of antiepileptic | One | 186 | 66.9 |
|  | More than one | 92 | 33.1 |
| Seizure attacks in the last month | None | 173 | 62.2 |
|  | One or more | 105 | 37.8 |
| Family history of epilepsy | No | 265 | 95.3 |
|  | Yes | 13 | 4.7 |
| Family history of mental illness | No | 251 | 90.3 |
|  | Yes | 27 | 9.7 |
| Medical comorbidities | No | 245 | 88.1 |
|  | Yes | 33 | 11.9 |

### Comorbid medical conditions

11.9% (n=33) of the respondents had comorbid medical illnesses. Hypertension was the most prevalent comorbid disease at 15.2% (n=5), followed by HIV, diabetes, gastritis, back pain and arthritis, each at 9.1% (n=3). These comorbid conditions are illustrated in Fig 1.

**Fig 1.**
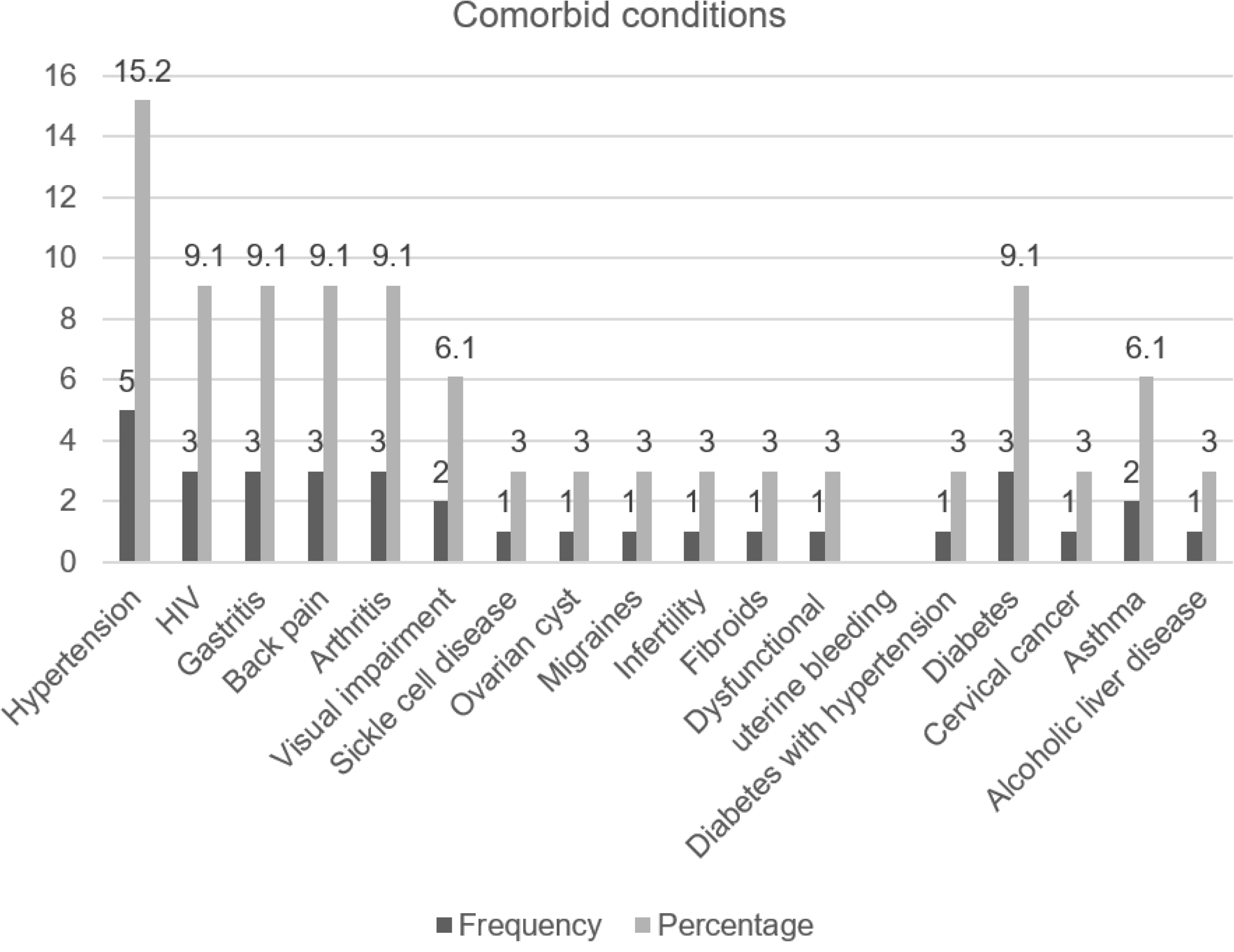
Comorbid medical conditions among the study participants. Bar chart showing the frequencies and percentages of different comorbid medical conditions among the study participants.

### Prevalence of psychiatric disorders

Of the respondents, 52.2% (n=145) had at least one psychiatric condition (95% CI: 95 % CI:46.3-58.0). Major depressive disorder was the most prevalent at 36.0%, followed by anxiety disorder cumulatively at 26.2% (generalized anxiety disorder at 16.9%, social anxiety disorder at 5.0%, agoraphobia at 3.2% and panic disorder at 1.1%). Psychotic disorder was present in 16.9%, while 15.1% respondents reported suicidality. Participants also presented with other psychiatric conditions as illustrated in Table 3.

**Table 3.** Prevalence of specific psychiatric disorders.

| Psychiatric disorder | Frequency (n) | Percentage (%) |
| --- | --- | --- |
| Major depressive episode | 100 | 36 |
| Generalized anxiety disorder | 47 | 16.9 |
| Social anxiety disorder | 14 | 5 |
| Agoraphobia | 9 | 3.2 |
| Panic disorder | 3 | 1.1 |
| Psychotic disorder | 47 | 16.9 |
| Suicidality | 42 | 15.1 |
| Alcohol use disorder | 25 | 9 |
| Other substance use disorder | 24 | 8.6 |
| Post-traumatic stress disorder | 14 | 5 |
| Manic episode | 18 | 6.5 |
| Hypomanic episode | 18 | 6.5 |
| Mood disorder with psychotic features | 8 | 2.9 |

**Table 4.** Determinants of psychiatric morbidity.

| Variable | Category | uOR | P-value | 95% CI | aOR | 95% CI | P-value |
| --- | --- | --- | --- | --- | --- | --- | --- |
| Gender | Male | Ref |  |  | Ref |  |  |
|  | Female | 1.80 | 0.029 | 1.05-2.73 | 1.602 | (0.831-3.087) | 0.59 |
| Marital status | Married | Ref |  |  | Ref |  |  |
|  | Single | 1.46 | 0.164 | 0.86-2.48 | 1.81 | (0.777-4.220) | 0.169 |
|  | Separated/ Widowed | 5.91 | 0.001 | 2.04-17.14 | 2.74 | (0.661-11.363) | 0.164 |
| Education level | Primary | Ref |  |  | Ref |  |  |
|  | Secondary | 0.10 | <0.001 | 0.03-0.30 | 0.302 | (0.086-1.062) | 0.062 |
|  | Tertiary | 0.08 | <0.001 | 0.03-0.25 | **0.221** | **(0.054-0.903)** | **0.036** |
| Employment status | Formally employed | 0.57 | 0.156 | 0.26-1.24 | 1.179 | (0.346-4.015) | 0.792 |
|  | Casually/Self employed | 1.73 | 0.035 | 1.04-2.89 | **2.590** | **(1.160-5.782)** | **0.020** |
|  | Unemployed | Ref |  |  | Ref |  |  |
| Duration of epilepsy | 1-10 years | Ref |  |  | Ref |  |  |
|  | ≥11 years | 3.84 | <0.001 | 1.92-7.70 | 1.355 | (0.111-1.139) | 0.082 |
| Seizure related trauma | No | Ref |  |  | Ref |  |  |
|  | Yes | 10.94 | <0.001 | 5.45-21.98 | **4.032** | **(1.552-10.476)** | **0.004** |
| No. of antiepileptics | One | Ref |  |  | Ref |  |  |
|  | More than one | 12.41 | <0.001 | 6.30-24.43 | **4.280** | **(1.790-10.230)** | **0.001** |
| Seizures in last month | None | Ref |  |  | Ref |  |  |
|  | One or more | 6.75 | <0.001 | 3.84-11.87 | **3.801** | **(1.875-7.704)** | **<0.001** |
| Family history of mental illness | No | Ref |  |  | Ref |  |  |
|  | Yes | 28.84 | 0.001 | 3.85-215.81 | 6.095 | (0.495-75.112) | 0.158 |
| Family history of epilepsy | No | Ref |  |  | Ref |  |  |
|  | Yes | 26.63 | 0.001 | 5.27-110.93 | 5.018 | (0.326-66.629) | 0.149 |
| Medical comorbidities | No | Ref |  |  | Ref |  |  |
|  | Yes | 6.13 | <0.001 | 2.29-16.39 | **5.478** | **(1.377-11.353)** | **0.047** |

Abbreviations: uOR, unadjusted odds ratio; aOR, adjusted odds ratio; CI, confidence interval

### Significant determinants of psychiatric morbidity

Participants with tertiary level of education had a nearly 80% reduced odds of psychiatric morbidity as compared to those with only primary level of education (aOR=0.221; 95% CI: 0.054–0.903; p=0.036). Being casually/self-employed had a nearly three times increased odds of psychiatric morbidity compared to being unemployed (aOR=2.590; 95% CI: 1.160–5.782; p=0.020). Among the clinical variables, having seizure-related trauma was associated with four times increased odds of psychiatric morbidity (aOR=4.032; 95% CI: 1.552–10.476; p=0.004).

Similarly, antiepileptic polytherapy also had four times increased odds of psychiatric morbidity compared to antiepileptic monotherapy (aOR=4.280; 95% CI: 1.790–10.230; p=0.001). Participants with recent seizure attacks had a nearly four times higher odds of developing psychiatric morbidity (aOR=3.801; 95% CI: 1.875–7.704; p<0.001), and those with comorbid medical conditions had five times increased odds (aOR=5.478; 95% CI: 1.377–11.353; p=0.047). Factors like gender, marital status, duration of epilepsy and a positive family history of epilepsy or mental illness were not significant predictors of psychiatric morbidity as shown in Table 6.

## Discussion

This study revealed a significant burden of psychiatric morbidity in persons living with epilepsy (PLWE), with 52.2% of the respondents presenting with one or more psychiatric diagnoses. This supports evidence that epilepsy is not simply a neurological disorder, but a neuropsychiatric disease with significant mental health effects [28,29]. The most common diagnoses were depression and anxiety disorders, which is consistent with previous findings of psychiatric morbidity in PLWE [30–39].

The high prevalence of psychiatric comorbidity in this population may be a result of many difficulties that are associated with living with a chronic neurological condition. Epilepsy not only impacts on the patient’s physical health but also on emotional, social and psychological functioning [40,41]. The constant uncertainty and fear associated with recurrent, unpredictable seizures can lead to significant stress and anxiety, which can take an emotional toll on patients [42–44]. The association of epilepsy and psychiatric comorbidity could also be due to stigma, societal exclusion, restrictions in the scope of academic and occupational pursuits, and worry about an uncertain future [43,44]. Biological factors may also play a role, as epilepsy and psychiatric disorders are thought to share common neurobiological mechanisms [33,43,45–47].

The overall prevalence of psychiatric morbidity in this study is similar to results from research done in India, where rates among PLWE have been estimated to be 50-52% [48,49]. It is however less than that found in a number of studies conducted in Sub-Saharan Africa, including those from Cameroon and Kenya [50,51]. These discrepancies could partially be explained by differences in the study sites and assessment tools. Psychiatric clinics typically review patients with severe psychiatric symptoms through referral systems and research conducted in such a setting, as was observed in the Kenyan study [51], is more likely to reveal a higher prevalence. In contrast to the structured diagnostic tools like the MINI, which was used in the current study and offers more precise psychiatric diagnoses, the screening tools frequently produce broader prevalence estimates, which may account for the findings in the Cameroonian study [50].

Depression was the most common mental condition found in this study. The common co-occurrence may result from the burden of emotional stress associated with epilepsy. Having repeated seizures, the anticipation of their occurrence, and the chronicity of the illness may lead to development of feelings of hopeless, sadness and loss of control [43]. Individuals living with epilepsy may also experience stigma, isolation and have difficulties in reaching goals in respect to life achievement in education, occupation, relationships and family, all of which may lead to depressive symptoms [43,44]. The co-occurrence can also be due to the shared neurobiological pathways between the two conditions [33,43], as well as the neuropsychiatric effects of some antiepileptic medications [44,52,53].

The prevalence of depression in this study is in line with research from Ethiopia and Nepal, as well pooled meta-analyses that show depression prevalence in more than one-third of patients living with epilepsy [54–57]. The use of highly sensitive screening tools, variations in illness severity, and possible referral bias may all contribute to the prevalence estimates found in research from Kenya and Burkina Faso which reported higher rates [58,59]. Conversely, reduced prevalence found in some Asian populations and private healthcare settings might be the result of variations in research demographics, healthcare accessibility and socioeconomic circumstances [60,61].

Anxiety disorder was the second most common psychiatric condition. Given the unpredictable nature of seizures, patients are likely to have constant fear and worry about when or where the next seizure may occur, and the possible consequences including injury, embarrassment or loss of independence [42–44]. Anxiety could also result from common neurobiological vulnerabilities underlying epilepsy and anxiety, particularly within the limbic system [33,47,62]. Some antiepileptic medications may also contribute to or worsen anxiety symptoms [44,53].

The anxiety prevalence in this study is consistent with international pooled estimates of 25%–26% [63,64], and results from research done in nations like Saudi Arabia, Turkey, and the United Arab Emirates [31,37,65]. Nearly one in six participants had experienced psychotic disorders, which was higher than estimates from other parts of the world [66,67], probably due to variances in patient demographics and methodological differences. Suicidality was found in about one in seven participants, lower than pooled global estimates [68], but consistent with findings from Ethiopian and earlier Kenyan studies [51,69]. This decreased prevalence may possibly be due to underreporting emanating from social desirability bias associated with stigma in low and middle-income countries, leading to an under detection rather than the true prevalence.

There was a substantial correlation between psychiatric morbidity and both clinical and sociodemographic variables. Participants with tertiary education had a nearly 80% reduced odds of psychiatric morbidity compared to those with only primary education (aOR = 0.221, 95% CI: 0.054–0.903, p = 0.036), indicating a strong protective effect of educational level. Access to healthcare services, coping strategies and health literacy may all be enhanced by higher educational attainment. Psychiatric illness was also substantially correlated with employment status.

Compared to participants who were unemployed, those who had casual/self-employment had higher odds of psychiatric morbidity (aOR = 2.590, 95% CI: 1.160–5.782, p = 0.020), perhaps as a result of financial and occupational insecurity or instability. On the other hand, students accounted for the majority (n=76, 63%) of the unemployed group. This may account for the lower psychiatric morbidity among the unemployed participants, as students were likely to be supported by their caregivers.

Among the clinical variables, seizure related injuries had a high correlation with psychiatric morbidity (aOR = 4.032, 95% CI: 1.552–10.476, p = 0.004), reflecting both the psychological effects of traumatic seizure events and the severity of epilepsy. Higher psychiatric morbidity was also linked to increased seizure frequency (aOR = 3.801, 95% CI: 1.875–7.704, p < 0.001), possibly because frequent and unexpected seizures impede functioning, create emotional distress and lower quality of life [27]. Another significant predictor of psychiatric morbidity was antiepileptic polytherapy (aOR = 4.280, 95% CI: 1.790–10.230, p = 0.001), which is suggestive of refractory epilepsy, increased treatment burden, and potential neuropsychiatric adverse effects. There was also a significant increase in the odds of psychiatric morbidity when additional medical comorbidities were present (aOR = 5.478, 95% CI: 1.377–11.353, p = 0.047). The accumulated burden of chronic illnesses, shared inflammatory pathways, and overlapping neurobiological mechanisms between mental disorders and physical illnesses could all contribute to this link.

These results are in line with earlier research showing that comorbid medical illnesses, antiepileptic polytherapy, and frequent seizures are significant contributors to poor mental health outcomes among patients living with epilepsy [20–22,24,25,70–72]. Overall, this study’s findings are consistent with a biopsychosocial model in which the severity of the neurological condition, treatment related variables, psychosocial stresses, and socioeconomic difficulties all work together to affect psychiatric morbidity of individuals living with epilepsy.

### Limitation of the study

Major limitations were the cross-sectional design which could only establish association but not causality, and limited generalizability of the findings considering that it was a hospital-based study, hence results are majorly applicable to similar tertiary settings. There was also a possibility of social desirability bias. Although participants were assured of confidentiality and interviews were conducted in private, the stigma attached to sensitive topics like suicidality could still result in participants withholding information which could lead to an underestimation of the true prevalence. Nevertheless, this study adds valuable information to inform integrated neuropsychiatric management in epilepsy care in this setting.

## Conclusion

This study revealed high rates of psychiatric morbidity among patients living with epilepsy, affecting more than half of the participants. The most common psychiatric diagnosis was depression, followed by anxiety, psychosis and suicidality. Frequent seizures, seizure-related physical trauma, antiepileptic polytherapy, comorbid medical conditions and non-formal employment were key predictors of psychiatric morbidity. Tertiary education on the other hand was a protective factor.

This study highlights the importance of routine screening for common mental health disorders in epilepsy treatment clinics, and the need to incorporate a multidisciplinary approach in epilepsy care. Practical interventions include better seizure control, regular medication review and optimization of treatment, with increased psychosocial support. Future research should include longitudinal studies to establish causality, and community-based studies to estimate the burden of psychiatric morbidity at the community level.

## Data Availability

The data supporting the findings of this study are available within the manuscript and its Supporting Information files. The sociodemographic/clinical characteristics questionnaire and the dataset have been included as Supporting Information.

## Acknowledgements

We express our appreciation to the administration and staff at Moi Teaching and Referral Hospital, Neurology clinic, for their support in the course of data collection. We also acknowledge the statistician who analyzed the data, and are indebted to the study participants for their participation in the study.

## Author contributions

**Conceptualization:** Agnes Achieng Odhiambo, Daniel Waiganjo.C Kinyanjui, Robina Kerubo Momanyi

**Data curation:** Agnes Achieng Odhiambo, Henry Mwangi

**Formal analysis:** Henry Mwangi

**Investigation:** Agnes Achieng Odhiambo

**Methodology:** Agnes Achieng Odhiambo, Daniel Waiganjo.C Kinyanjui, Robina Kerubo Momanyi

**Project administration:** Agnes Achieng Odhiambo

**Resources:** Agnes Achieng Odhiambo

**Supervision:** Daniel Waiganjo.C Kinyanjui, Robina Kerubo Momanyi

**Validation:** Agnes Achieng Odhiambo, Daniel Waiganjo.C Kinyanjui, Robina Kerubo Momanyi

**Visualization:** Agnes Achieng Odhiambo, Henry Mwangi

**Writing-original draft:** Agnes Achieng Odhiambo

**Writing-review & editing:** Agnes Achieng Odhiambo, Daniel Waiganjo.C Kinyanjui, Robina Kerubo Momanyi, Henry Mwangi

## Supporting information

**S1 File. Sociodemographic and clinical characteristics questionnaire.**

Researcher designed questionnaire used for collecting sociodemographic and clinical data.

**S2 File. Study dataset.** Excel workbook containing de-identified dataset.

**S3 File. Ethical approval letter.** Approval letter granted by the Moi University/Moi Teaching and Referral Hospital Institutional Research and Ethics Committee (IREC).

**S4 File. NACOSTI research authorization permit.** Research permit granted by the National Commission for Service, Technology and Innovation (NACOSTI).

